# Educational expectations and needs of individuals scheduled for breast surgery within the Primary Nursing Model: a qualitative study

**DOI:** 10.64898/2026.10.25.26365062

**Authors:** Mariateresa Sciscioli, Andrea Moro, Stefania Grisanti, Giorgio Magon, Duilio Fiorenzo Manara

**Affiliations:** European Institute of Oncology, IRCCS, Milan, Italy; Master’s Degree in Nursing and Midwifery Sciences Università Vita-Salute San Raffaele, Milan, Italy; Center for Nursing Research and Innovation (CeNRI) Università Vita-Salute San Raffaele

**Keywords:** Breast Surgery, Nurse-Led Patient Education, Primary Nursing

## Abstract

**Background:** A diagnosis of breast cancer may represent a potentially destabilizing event, with significant physical, psychological, and social implications. Individuals scheduled for breast surgery require clear, understandable, and personalized information to prepare for surgery and manage the postoperative period. However, a discrepancy may arise between perceived informational needs and the information actually received. The Primary Nursing model, based on continuity of care and the presence of a designated primary nurse, may promote greater personalization of nurse-led patient education.

**Aim:** To explore the experiences of individuals scheduled for breast surgery in relation to the nurse-led patient education received throughout the perioperative pathway, in order to understand their educational expectations, informational needs, and perceptions of the usefulness of the educational interventions provided within the Primary Nursing model.

**Design:** A qualitative study with a phenomenological orientation.

**Methods:** The study will be conducted at the European Institute of Oncology. Participants will be recruited through convenience sampling among individuals diagnosed with breast cancer who have undergone surgery and received the nurse-led discharge education routinely provided. Approximately 20 participants are expected to be enrolled; the final sample size will be determined according to the principle of data saturation. Data will be collected through semi-structured interviews conducted approximately two weeks after discharge by a member of the research team who is not involved in the participants’ clinical care. The interviews, lasting approximately 30–40 minutes, will be audio-recorded with prior consent, transcribed verbatim, and assigned a sequential identification code. Sociodemographic data and clinical information extracted from the medical records will also be collected. The transcripts will be analyzed using thematic analysis according to Braun and Clarke, with a phenomenological orientation.

**Expected contribution:** The study may contribute to understanding potential discrepancies between educational expectations, informational needs, and the perceived usefulness of the information received, thereby informing clinical practice toward personalized educational interventions consistent with continuity of care.

## Background

A diagnosis of breast cancer represents a potentially destabilizing event, abruptly initiating a complex treatment pathway characterized by multiple clinical decisions and substantial physical, psychological, and social implications. In this context, the need for clear, understandable, and personalized information emerges from the earliest stages of care. The indication for breast surgery, often experienced as a pivotal and highly significant point in the treatment pathway, entails bodily, functional, and emotional changes that require adequate preparation not only from a clinical perspective, but also from educational and relational perspectives.

Several studies have shown that structured preoperative educational interventions can significantly improve the psychological preparedness of individuals scheduled for breast surgery by promoting a better understanding of the surgical pathway and contributing to a more positive overall experience of hospitalization and postoperative recovery. (İlgin et al. 2024) demonstrated that targeted educational programs can reduce anxiety and uncertainty among individuals awaiting surgery, while enhancing their sense of control and preparedness for the surgical pathway. Similarly, a qualitative meta-synthesis (Carr et al. 2019) found that many individuals report specific informational needs concerning the surgical procedure, the possibility of breast reconstruction, and the psychophysical consequences of surgery, while emphasizing that such information is not always adequately provided or tailored to individual needs.

A frequently reported issue in the literature is the discrepancy between individuals’ perceived informational needs and the information actually received throughout the care pathway. Certain topics, such as surgical drain management, upper-limb mobilization, pain management, and surgical wound care, may be perceived as insufficiently addressed, whereas other information may be considered redundant or poorly aligned with the individual’s actual needs immediately before or after surgery. This informational mismatch may adversely affect the individual’s ability to cope with the postoperative period and independently manage the transition home, thereby influencing the overall quality of the care experience.

Within this context, self-care plays a central role in the care pathways of individuals undergoing breast surgery. The individual’s ability to manage symptoms, dressings, functional limitations, and bodily changes depends largely on the quality and clarity of the education received during hospitalization. Recent studies confirm that nursing support focused on self-care and based on progressive, practical, and personalized educational interventions promotes the development of postoperative self-management skills, reduces the risk of preventable complications, and increases the individual’s sense of safety at discharge (Solera-Gómez et al., 2022). In particular, education on surgical drain management, recognition of signs of infection, pain management, and the gradual resumption of activities of daily living is a fundamental component of nursing care in oncological surgery units.

In addition to the quality of the information provided, the organizational model of care may significantly influence patients’ educational experience. The Primary Nursing model, adopted at the European Institute of Oncology (IEO) to ensure continuity of care and personalized care delivery, is based on the presence of a designated primary nurse who is responsible for planning and coordinating care throughout hospitalization. This model fosters a more stable and continuous care relationship between the nurse and the individual receiving care, thereby creating the conditions for an educational process that is more closely tailored to individual needs. The literature has shown that continuity-based care models may improve the consistency of the information provided, promote greater personalization of educational interventions, and increase patient satisfaction (Cocchieri et al., 2021; Gonçalves et al., 2023).

Despite the extensive literature on educational interventions in oncological surgery, there is still limited understanding of how individuals scheduled for breast surgery actually experience the education received during hospitalization and the extent to which that experience meets their informational needs and supports preparation for self-care after discharge. Furthermore, the role of the nursing organizational model in shaping the effectiveness of patient education remains underexplored.

In particular, the Primary Nursing model, which is based on continuity of care and the presence of a designated primary nurse, may promote greater personalization of nurse-led patient education and closer alignment with the informational needs of individuals scheduled for surgery.

Given these considerations, an in-depth exploration of the experiences of individuals scheduled for breast surgery in relation to the nurse-led patient education received throughout the perioperative pathway is warranted, in order to understand their educational expectations, informational needs, and perceptions of the usefulness of the information provided. A better understanding of these aspects may help guide clinical practice toward educational interventions that are more personalized and more consistent with the principles of continuity of care, while strengthening the nurse’s role in supporting individuals as they adapt to illness and independently manage the postoperative period.

### Research question

What is the lived experience of individuals scheduled for breast surgery regarding the nurse-led patient education received throughout the perioperative pathway within the Primary Nursing care model?

### Primary objective

The main aim is to explore the experiences of individuals scheduled to undergo breast surgery in relation to the nurse-led patient education they receive throughout the perioperative pathway, in order to understand their educational expectations and their perceptions of the usefulness of the information and support provided within the Primary Nursing care model.

### Secondary objectives

- To explore individuals’ educational expectations at the time of breast cancer diagnosis and before surgery.
- To describe individuals’ educational needs throughout the perioperative pathway.
- To understand how individuals interpret and assign meaning to the educational interventions received during hospitalization.
- To describe which educational topics are perceived as particularly useful or, conversely, as poorly relevant or insufficient for managing the transition home and the postoperative period.
- To explore how the educational relationship with nursing staff and the continuity of care ensured by the Primary Nursing model influence perceptions of the education received.

### Study inclusion criteria

The study will include all individuals who:

- have been diagnosed with breast cancer and are scheduled to undergo quadrantectomy with or without breast reshaping, mastectomy with or without breast reconstruction, and with or without axillary lymph node dissection;
- are admitted under the Italian National Health Service (Servizio Sanitario Nazionale, SSN) and/or as self-paying patients;
- have completed surgical treatment at the IEO and have received the nurse-led discharge education routinely provided;
- are able to understand and describe their educational experience;
- have read, understood, and signed the participant information sheet and the informed consent form for participation in the study.

The study will exclude individuals who:

- are undergoing neoadjuvant therapy before surgery or have previously undergone breast surgery;
- have been diagnosed with breast cancer and are scheduled to undergo surgery in a day-surgery setting;
- have clinical or cognitive conditions that preclude informed participation;
- do not provide informed consent to participate.

### Study design

To achieve the proposed objectives, a qualitative study with a phenomenological orientation will be conducted.

### Sample

Participants will be recruited through convenience sampling among IEO patients who have been diagnosed with breast cancer, have undergone breast surgery, and have received the nurse-led discharge education routinely provided as part of the care pathway.

Approximately 20 participants are expected to be enrolled, with potential participants identified with the support of the Breast Nurse. The final number of participants will nevertheless be determined according to the principle of data saturation, as recommended for qualitative studies with a phenomenological orientation.

Eligible individuals will be informed about the study by a member of the research team who is not directly involved in their care pathway. It will be made clear that participation is voluntary and that refusal to participate or withdrawal from the study will have no consequences for their care.

## Materials and methods

In order to address the research question, a qualitative design with a phenomenological orientation was selected to explore the lived experiences of enrolled participants in relation to the nurse-led patient education received throughout the perioperative pathway. Data will be collected through semi-structured interviews and a sociodemographic questionnaire. The interviews will explore both participants’ initial educational expectations and their retrospective appraisal of the educational experience after returning home, thereby enabling the identification of any discrepancies between anticipated informational needs and the perceived usefulness of the information received.

The interviews will be conducted by a member of the research team who is not directly involved in the participants’ clinical care, with the aim of minimizing the relational bias. Each interview will last approximately 30–40 minutes. Participants may stop the interview at any time should they experience emotional distress or discomfort.

The interviews may be conducted:

- in person, in a private setting at the European Institute of Oncology;
- or remotely through the institutional Microsoft Teams platform, if necessary.

All interviews will be audio-recorded with participants’ prior consent, transcribed verbatim in full, and assigned a sequential identification number before analysis.

### Data collection phases

The data collection procedure comprises several sequential phases designed to ensure transparent recruitment, informed participation, and that the interview is conducted at the most appropriate time for each individual.

During the preadmission phase, with the support of the Breast Nurse, eligible individuals will be invited to participate in the study. At that time, they will receive clear information regarding the study objectives, interview procedures, participants’ rights, and the management of personal data. Individuals will be asked to read and sign the informed consent form for study participation and data processing. Adequate time will be provided to read and understand the information, thereby supporting a free and informed decision.

Subsequently, within the first 24 hours after surgery, a member of the research team who is not directly involved in the care pathway will meet with the enrolled individual to reconfirm their willingness to participate, verify their understanding of the study, and jointly agree on the interview format and timing. This procedure will make it possible to respect the individual’s clinical and emotional needs during the postoperative period while ensuring the methodological appropriateness of the data collection process.

The interview will be conducted approximately two weeks after discharge to explore how participants experienced the education received throughout the perioperative pathway and the extent to which their initial educational expectations were met.

The interview guide will include questions such as:

1. *Could you tell me what educational support you expected to receive from the nursing staff at the time of diagnosis and before surgery?*
2. *Thinking back to what you expected before surgery, could you tell me how the education provided by your primary nurse did or did not meet your needs?*
3. *Could you tell me which educational instructions or explanations were most helpful when you returned home?*
4. *Looking back on your pathway—from preparation for discharge through managing at home—could you describe your experience of the information, explanations, and instructions you received? Which aspects were most useful? Were there any aspects you would have liked to have clarified, explored in greater depth, or received differently? Looking back, is there anything that would have been helpful to know before discharge?*
5. *Could you tell me whether there were any concerns, fears, or difficulties for which you would have liked more educational support from the nursing staff?*
6. *The primary nurse is the designated nurse who supports the individual throughout the care pathway, from admission to discharge, ensuring continuity and educational support. How would you describe your relationship with your primary nurse during your care pathway?*

### Data analysis

Interview transcripts will be analyzed using thematic analysis (Braun & Clarke, 2021) with a phenomenological orientation, with the aim of identifying recurrent themes and meanings within participants’ educational experiences.

The analysis will proceed through the following phases:

1. Repeated reading of the transcripts to become familiar with the data;
2. Initial coding of relevant content;
3. Identification of emerging categories and themes;
4. Review and definition of the main themes;
5. Interpretation of the findings.

Data collection will continue until data saturation is reached, that is, until the interviews no longer generate new relevant information.

In addition, study participants will be asked to complete a sociodemographic form designed to collect personal and sociodemographic data, information on the surgical procedure received, and other relevant clinical data extracted from the medical record. The sociodemographic questionnaire includes the following items:

- Age (years):________________
- Sex
  ▪ Female
  ▪ Male
  ▪ Prefer not to answer
- Region of residence
  ▪ Abruzzo
  ▪ Basilicata
  ▪ Calabria
  ▪ Campania
  ▪ Emilia-Romagna
  ▪ Friuli Venezia Giulia
  ▪ Lazio
  ▪ Liguria
  ▪ Lombardy
  ▪ Marche
  ▪ Molise
  ▪ Piedmont
  ▪ Apulia
  ▪ Sardinia
  ▪ Sicily
  ▪ Tuscany
  ▪ Umbria
  ▪ Aosta Valley
  ▪ Veneto
  ▪ Autonomous Province of Trento
  ▪ Autonomous Province of Bolzano
  ▪ I reside abroad
- **Marital status**
  ▪ Single
  ▪ Married / in a civil union / cohabiting
  ▪ Widowed
  ▪ Prefer not to answer
- **Employment status**
  ▪ Employee
  ▪ Not employed
  ▪ Self-employed professional
  ▪ Retired
  ▪ Not seeking employment
  ▪ Unemployed
  ▪ Other
  ▪ Student
- Do you have a caregiver or one or more other people supporting you throughout this pathway?
  ▪ Yes
  ▪ No
- Type of surgical procedure performed
  ▪ _______________________________________________________
- Did you undergo breast reconstruction with an implant, tissue expander, or flap?
  ▪ Yes
  ▪ No
- Were surgical drains still in place at discharge:
  ▪ Yes
  ▪ No

Information on existing comorbidities, any postoperative complications, and the surgical procedure performed will be retrieved from the medical record.

The information collected will be used exclusively to describe the sample and support the interpretation of the qualitative data.

### Expected study duration

Twelve months from approval by the Ethics Committee.

## Data Availability

All data produced in the present study are available upon reasonable request to the authors

## Data Availability

All data produced in the present study are available upon reasonable request to the authors

## Data confidentiality

Pursuant to Regulation (EU) 2016/679 of the European Parliament and of the Council of 27 April 2016 on the protection of natural persons with regard to the processing of personal data, participants are informed that personal data will be collected and stored electronically and used exclusively for scientific research purposes. Access to these data will be restricted and secured by the investigator. By signing the informed consent form, enrolled participants will authorize access to these data. The study results may be published.

### Informed consent

For research purposes, informed consent to participate and consent to the processing of personal data will be obtained from all individuals involved in the study in accordance with applicable data-protection legislation. All data will be coded and processed in anonymized form to protect the privacy of enrolled participants throughout all stages of the study.

### Data management and document retention

All audio and video recordings of the interviews, anonymized transcripts, and informed consent forms will be retained for five years.

### Use of information and publication of results

The data collected will be used to prepare the degree thesis of one member of the research team and will be disseminated to the scientific community through participation in conferences on the topic and through potential scientific publications.

## References

Braun, V., & Clarke, V. (2021). Thematic analysis: A practical guide. Sage.

Carr, T. L., Groot, G., Cochran, D., & Holtslander, L. (2019). Patient Information Needs and Breast Reconstruction After Mastectomy: A Qualitative Meta-Synthesis. Cancer Nursing, 42(3), 229–241. 10.1097/NCC.0000000000000599

Cocchieri, A., Magon, G., Cavalletti, M., Cristofori, E., & Zega, M. (2021). Exploring hospital compliance with the primary nursing care model: Validating an inventory using the Delphi method. BMC Nursing, 20(1), 188. 10.1186/s12912-021-00712-1

Gonçalves, I., Mendes, D. A., Caldeira, S., Jesus, É., & Nunes, E. (2023). The Primary Nursing Care Model and Inpatients’ Nursing-Sensitive Outcomes: A Systematic Review and Narrative Synthesis of Quantitative Studies. International Journal of Environmental Research and Public Health, 20(3), 2391. 10.3390/ijerph20032391

Ilgin, V. E., Yayla, A., & Kılınç, T. (2024). The Effect of Preoperative Education Given to Patients Who Will Have a Mastectomy: A Randomized Controlled Trial. Journal of PeriAnesthesia Nursing, 39(1), e1–e8. 10.1016/j.jopan.2023.09.007

Solera-Gómez, S., Benedito-Monleón, A., LLinares-Insa, L. I., Sancho-Cantus, D., & Navarro-Illana, E. (2022). Educational Needs in Oncology Nursing: A Scoping Review. Healthcare, 10(12), 2494. 10.3390/healthcare10122494

